# Human lipoproteins comprise at least 12 different classes that are lognormally distributed

**DOI:** 10.1101/2021.03.19.21253934

**Authors:** Tomokazu Konishi, Risako Fujiwara, Tadaaki Saito, Nozomi Satou, Naoko Crofts, Ikuko Iwasaki, Yoshihisa Abe, Shinpei Kawata, Tatsuya Ishikawa

## Abstract

Lipoproteins in medical samples have been measured by enzymatic methods that coincide with conventional ultracentrifugation. However, the high gravity and time required for ultracentrifugation can cause sample degradation. This study presents the results of HPLC, a gentler and rapid separation method, for 55 human serum samples. The elution patterns were analysed parametrically, and the attribute of each class was confirmed biochemically. Human samples contained 12 classes of lipoproteins, each of which may consist primarily of proteins. There are three classes of VLDLs. The level of each class was distributed lognormally, and the standard amount and the 95% range were estimated. Enzymatic methods measure the levels of several mixed classes. This lognormal character suggests that the levels are controlled by the synergy of multiple factors.

## Introduction

Lipoproteins are measured in two ways. The first is related to class separation for biochemical purposes ^1-4^. This method uses ultracentrifugation, which spontaneously creates a salt density gradient due to centrifugal forces. Although the classes of lipoproteins are separated according to their density, this method is time-consuming and is not suitable for measuring large numbers of medical samples. The other method, which uses enzymatic processes, is used for physical examination ^5^. In this method, cholesterol is chemically extracted from a certain class of lipoproteins and then measured. Several kits are available, but all are adjusted to mimic the results of the ultracentrifugation method. In the ultracentrifugation approach, the larger the particle size, the lower the density of lipoproteins; therefore, names such as very low-density lipoprotein (VLDL) and high-density lipoprotein (HDL) are used for large and small particles, respectively ^6,7^.

The accuracy of ultracentrifugation has been questioned in a study using rat serum ^8^. The very high centrifugal force was sufficient to pull hydrophobic proteins out of the membrane ^9^; additionally, complete separation can take several days, during which proteins can be degraded. There is a gentler way to separate these classes via HPLC gel filtration. This takes up to 30 min and does not require extra salts ^10^. This is not a novel method, but there were a few issues with how the data were analysed, as it was noted that the data were intended to be consistent with standard ultracentrifugation results. Analysis of HPLC results of rat data with parametric analysis (Materials and Methods) showed striking differences from the ultracentrifugation results ^8^. All classes of lipoproteins are protein-rich particles, contrary to conventional knowledge ^6,7^. Two new classes, LDL-antiprotease complexes (LAC), were also discovered.

## Results and discussion

At least 12 classes of lipoproteins in normal distribution had to be presumed to fit the human data (Fig. 1); even a minor class cannot be ignored (Fig. S2). The attributes of the classes were determined based on the elution pattern of the major protein components (Fig. 1, S3-S6). The elution patterns of these proteins coincided with the distribution of the corresponding classes. As the positions and scales coincided between TG and cholesterol, it was confirmed that a class had a fixed ratio of these substances.

**Fig. 1.**
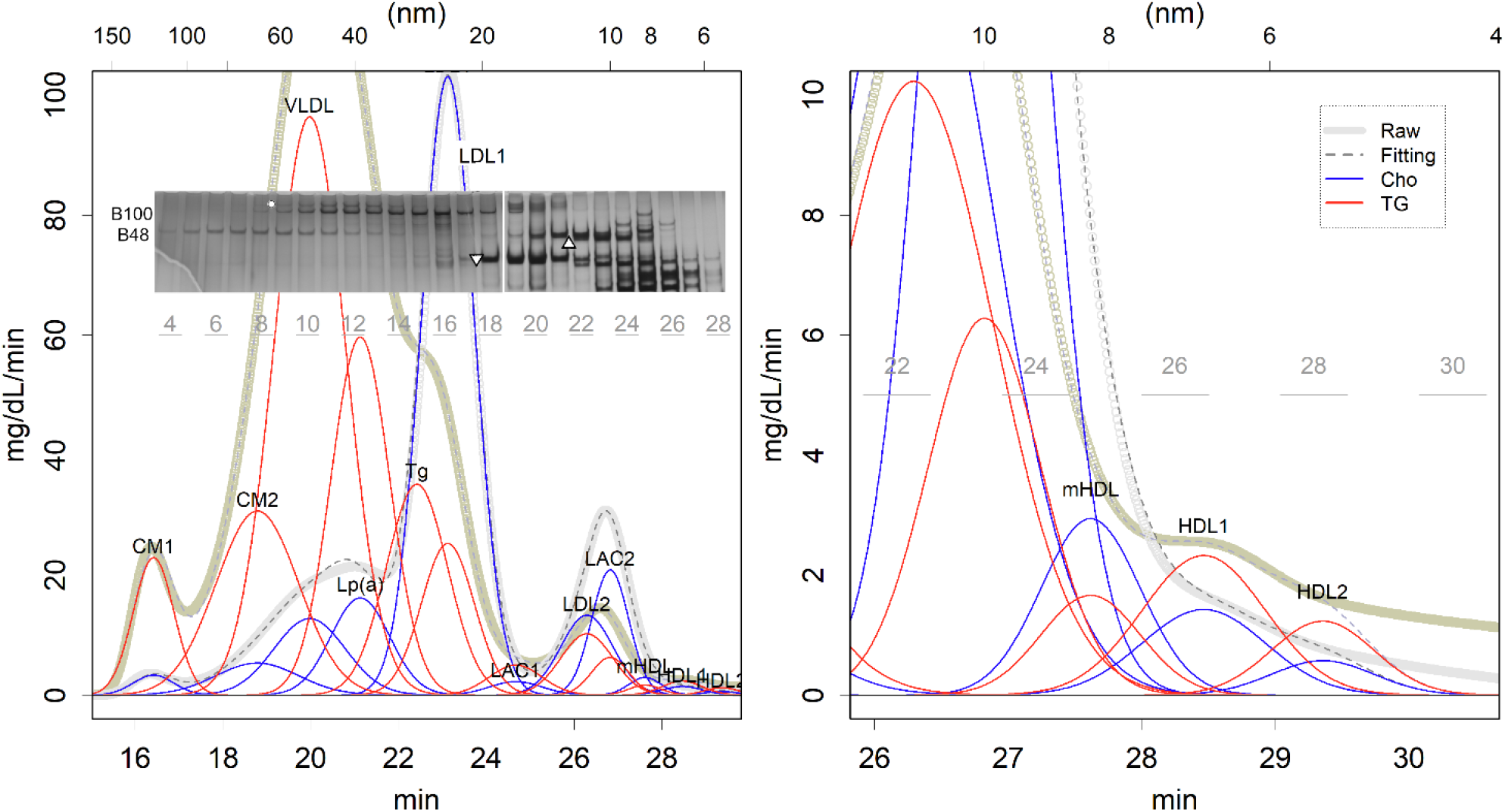
Elution pattern of HPLC of a hyperlipidaemia patient. In gel filtration, the shorter the elution time, the larger the particle size; elution time is proportional to the logarithms of the diameter of the particles. Left: whole image, right: enlarged view around HDLs. The bold line presents the measurement raw data, the red (TG) and blue (cholesterol) lines are the curve-fitted classes, and the dotted lines are their sum. There are 12 classes. The superimposed photo is a part of SDS-PAGE. B100 and B48 are the respective ApoB positions. ▿: Alpha-2-macroglobulin, ▵: Inter-alpha-trypsin inhibitor heavy chain, ○: Lp(a). Here an example of a hyperlipidaemic patient is shown so that the classes can be easily observed. An alternative to healthy volunteers is shown in S6 Fig.

Many features in human samples were similar to rat results ^8^. Each elution pattern was a mixture of normal distributions (Fig. 1 and S2-S6). This shows that each class of lipoprotein is stable in serum, which is inconsistent with the scenario in which CM and VLDL lose TG by gradual degradation ^6,7^, which in turn produces more skewed distributions ^8^. Rather, a single degradation of TG should have converted them into the next class at once.

CM and VLDL were eluted, consistent with observed peaks of ApoB48 and ApoB100, respectively (Fig. 1A and S3). Most ApoB proteins appear to be degraded during TG removal. In fact, the LDL1 fraction contains less B100 than expected for a large number of particles, and the LDL2 fraction is devoid of B48. HDLs were eluted with ApoA-1 (Fig. S4). Fractions of LAC1 and LAC2 eluted with major antiproteases, Alpha-2-macroglobulin and inter-alpha-trypsin inhibitor, respectively, similar to those in rats (Fig. S5) ^8^. Antiproteases, and thus LACs, are synthesised in the liver ^11,12^; they may supply cholesterol to the thrombus and protect it from lysis by plasmin.

There were some differences observed between rat and human lipoproteins. There were probably three classes of VLDL particles in human lipoproteins, which contained ApoB100 (Figs. 1 and S3), compared to rat lipoprotein which only has one class of VLDL particles. These lipoproteins may be secreted by the liver. Here, the largest class is denoted as VLDL. Another cholesterol-rich class included another lipoprotein denoted as Lp(a) protein ^13,14^ (Fig. 2A). The size of the particles seems to be larger than that previously reported ^13^; ultracentrifugation may have removed a portion of the particles. Another smaller class was noted to have a size slightly larger than that of LDL1. Estimating the cholesterol content of this class was difficult as its size is close to that of LDL1. However, as the fitting results suggested low cholesterol (S2 Table), this is denoted as the TG-rich class (TR) in this study. The levels of VLDL, Lp(a), and TR showed no relationship, suggesting that they were produced independently (Fig. S8AB). Rather, the cholesterol carrier, Lp(a), showed a weak negative correlation with the levels of another carrier, LDL2 (Fig. S8C).

**Fig. 2.**
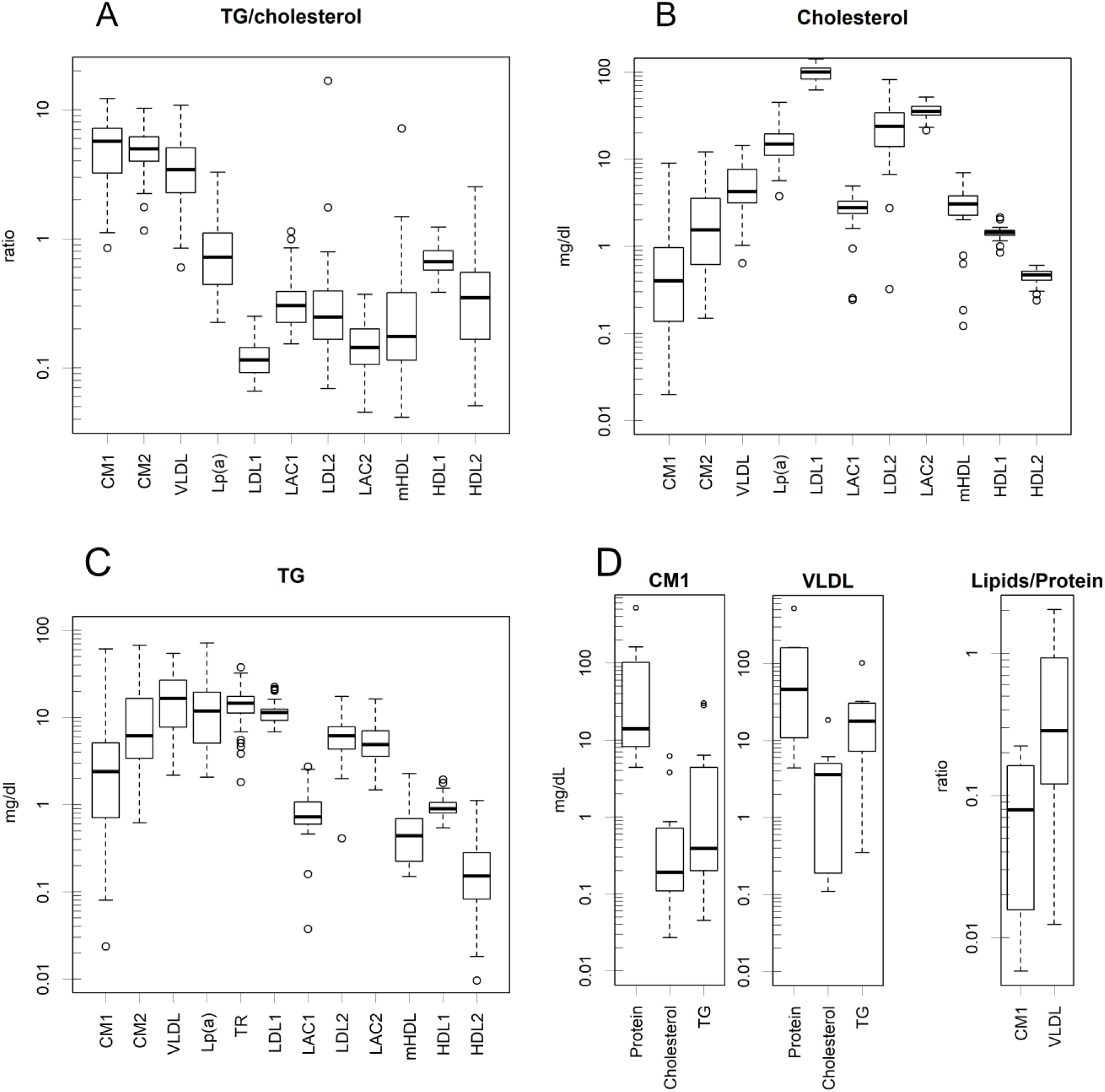
Box plot of the logarithm of quantity. All the y-axes are on a logarithmic scale. **A**. The ratio of TG/cholesterol. Naturally, the ones produced with TG are higher. The contents are **B**. Cholesterol and **C**. TG (mg / dL). While LDL1, LACs, and HDLs appeared within a certain narrow range, CM and VLDL levels fluctuated. **D**. Amounts and ratios of CM and VLDL measured by the preparative HPLC of healthy samples. Lipids (TG + cholesterol) are less than proteins in those classes.

There were two sizes of larger particles with the largest particles with B48 denoted as CM1. The 50-60 nm particles had B48 (Figs. 1, S3, and S6), and the level showed a higher correlation with CM1 than VLDL (Fig. S8DE); hence, the class is denoted as CM2. These particles would be intestine-derived ^6,7^.

The particle size and ApoB protein distribution suggest that they can be classified into three lineages:

1. CM → LDL2 → LAC2
2. VLDL, Lp(a) → LDL1 → LAC1
3. HDL2 → HDL1 → mHDL.

These protein distributions were the same as those of the rats ^8^. In this study, we focused on determining the attributes; hence, we did not analyse other apoproteins such as ApoC or ApoE. Certainly, we may only identify a fraction of apoproteins ^8^. Additionally, it should also be noted that the number of classes presented here was the smallest to perform curve fitting (Fig. S2). Systems with better separation may find more classes. The positions and scales of the classes are listed in Table S1. Classes that were initially secreted from the liver or intestine showed a larger scale. The particles may have a larger tolerance for the ratio and amount of lipids, which may have expanded this parameter. However, the elution position of the class did not change significantly between the samples. Therefore, the estimated peak diameters were within a certain range, indicating the physical stability of the particles (Table S1).

The amount of each class varied significantly among the volunteer samples (Fig. 2BC, C). They were lognormally distributed, as was found from the logarithms of the data quantiles, which showed a linear relationship with the theoretical values of the normal distribution. Linear correlations were confirmed in all classes (Fig. S7); The slope and intercept of the regression line represent σ and μ of the normal distribution, respectively; these are the distributions used to fit the curves of each sample. The parameters were estimated by robust methods, specifically MAD and trimmed means; the appropriateness of the estimation can be checked to determine whether the lines fit the plots. By accumulating z-normalized data using these parameters, *z*_i_ = (log(*x*_i_) -μ) / σ for any raw data *x*_i_, the distribution can be confirmed in a more exact quantile-quantile (QQ) plot (Fig. 3), where the slope is 1 and the intercept is zero. The lower part of the QQ plot bends downward, which is considered to be an artefact during curve fitting because low values are easy to ignore.

**Fig. 3.**
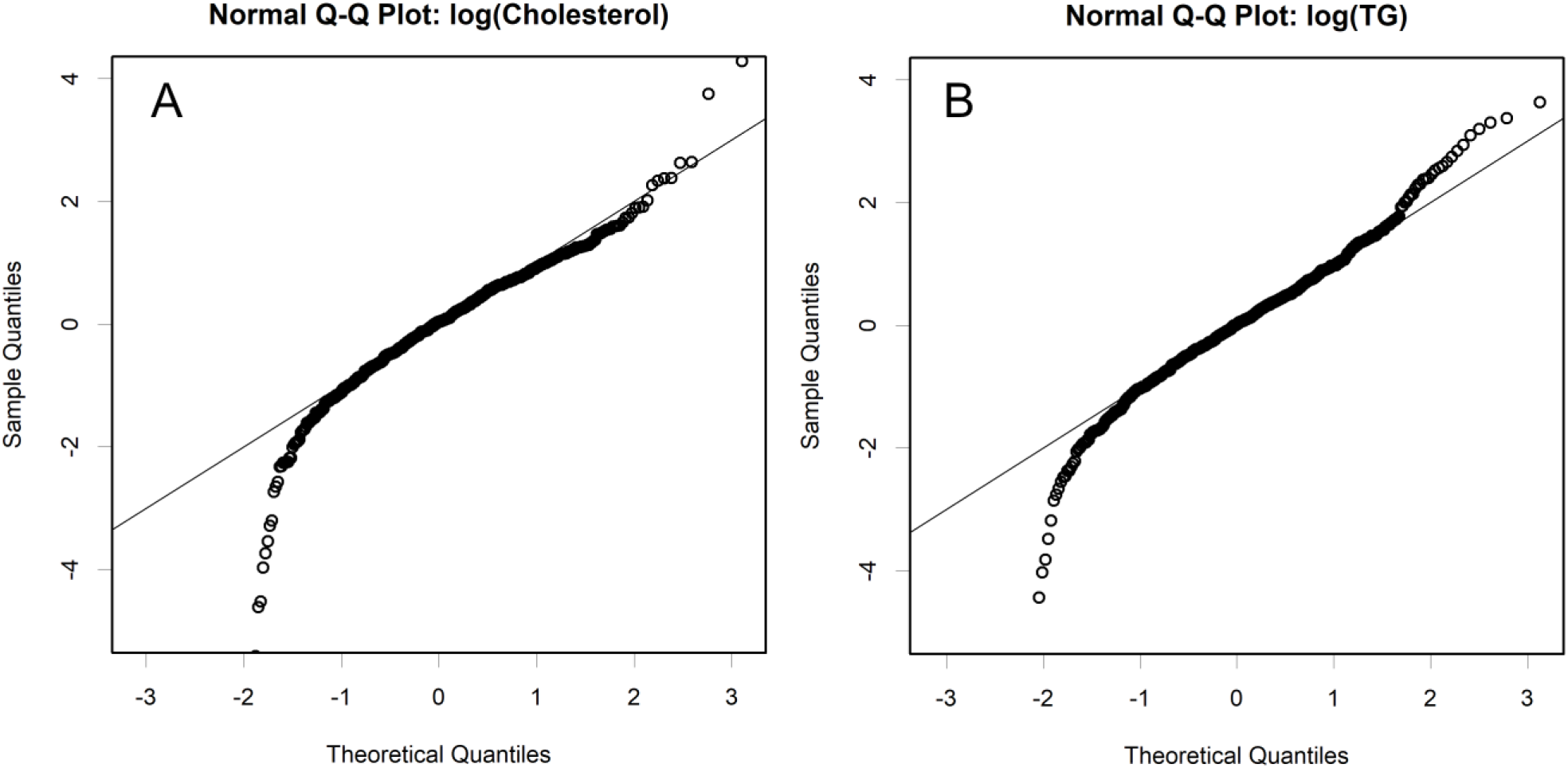
Normal QQ plot of the logarithms of normalized data. Cholesterol (**A**.) and TG (**B**). Quantiles are compared between the theoretical normal distribution and the z-normalized log-data. Here a lognormal distribution will form a straight relationship.

It is not surprising that each class was lognormally distributed. For example, the transcriptome has the same distribution properties ^15^. Levels of lipoproteins, as well as mRNA, are regulated by a balance between synthesis and degradation, both of which are controlled by specific factors in a multiplicative manner. This mechanism determines the distribution (Fig. S11). Therefore, lipoprotein levels can easily change due to multiple small causes; differences in multiple lifestyle habits may worsen the medical condition. In contrast, simultaneous clinical efforts may synergistically improve lipoprotein levels. This would explain why a different set of feeds drastically altered lipoprotein levels in a rat study ^16^.

Knowing that the data is lognormally distributed, the standard value and the 95% range of the classes could be estimated with accuracy, even though they were largely dispersed (Table S2). Note that the interval ranges inevitably become asymmetric to the standards: the higher is always wider (Fig. 11B).

The ratio of TG to cholesterol in each class also fluctuated greatly (Fig. 2A). Naturally, these ratios are large in the class synthesised to include TG, such as CM and VLDL. HDL1 and 2 are considered precursors to HDL (Fig. S4). They mature by receiving cholesterol.

LDL1 was the most predominant cholesterol carrier, followed by LAC2 (Fig. 2B). Some cholesterol carriers appeared within certain narrow ranges (LDL1, LACs, and HDLs). These classes may maintain cholesterol homeostasis. Conversely, variations in CM and VLDL levels were particularly high (Fig. 2B). The former may depend on diet, and the latter may depend on the body’s requirements for TG as a storable energy source. Among the TG-rich classes, TR appeared to be fairly stable.

HDLs were minor cholesterol carriers, contradicting the estimations of the previous study, which did not check whether each of the fractions contained ApoA-1 protein ^17^. In the study by Gordon et al., they assigned the whole of a major peak of cholesterol as HDL, which seemed to be a reasonable decision, since HDL was believed to be the major cholesterol carrier from ultracentrifugation results ^6,7^. However, in reality, the peak size is too large to be assigned to HDL alone; the structure of HDL is surrounded by ApoA-1 and therefore has a limitation in size ^18^. In fact, ApoA-1 was only confirmed at the end of the peak (Fig. S4). Rather, the peak was mainly composed of LDL2 and LAC2 (Fig. 1); these classes may have behaved as high-density particles during ultracentrifugation. However, their origins and functions are quite different from those of HDL.

In contrast to rats, human serum contained higher amounts of immunoglobulins. It should be noted that rats grown in pathogen-free environments did not show detectable levels of these proteins in SDS-PAGE ^8^. Some of them were as large as certain classes of lipoproteins (Fig. S9), which interfered with the estimation of protein abundance in these classes.

In contrast, CM and VLDL are very large particles; hence, the corresponding fractions would be free from immunoglobulins. The amount of protein was estimated from the UV record of preparative HPLC, and the ratios of the lipids (TG and cholesterol) were observed (Fig. 2D). The contents of both classes varied greatly; however, they always contained fewer lipids than protein, contradicting the results of ultracentrifugation ^6,7^.

The amounts of HDL and LDL measured by conventional enzymatic methods were much higher than those of any of the classes. This is not surprising, as those account for the majority of total cholesterol (Fig. S10). If conventional methods extract cholesterol from certain classes with high efficiency, the observed data would reflect combinations of some classes. Such combinations were estimated as combinations with the highest correlation and similarity (Fig. 4). The conventional measure of LDL would be VLDL derivatives without VLDL, and HDL would be HDLs and CM derivatives without CM1. Of course, we cannot deduce any of the true classes from the conventional measurements. In particular, the estimated HDL differed significantly from the actual amount of HDLs. Moreover, sums of lognormally distributed quantities are not informative at all, as an anomalous class may make up the majority of the sum; in a statistical sense, they do not even show average levels (Fig. S12). Unfortunately, it is also true that total TG or cholesterol does not provide useful information. Rather, an exact measurement of each class is desirable.

**Fig. 4.**
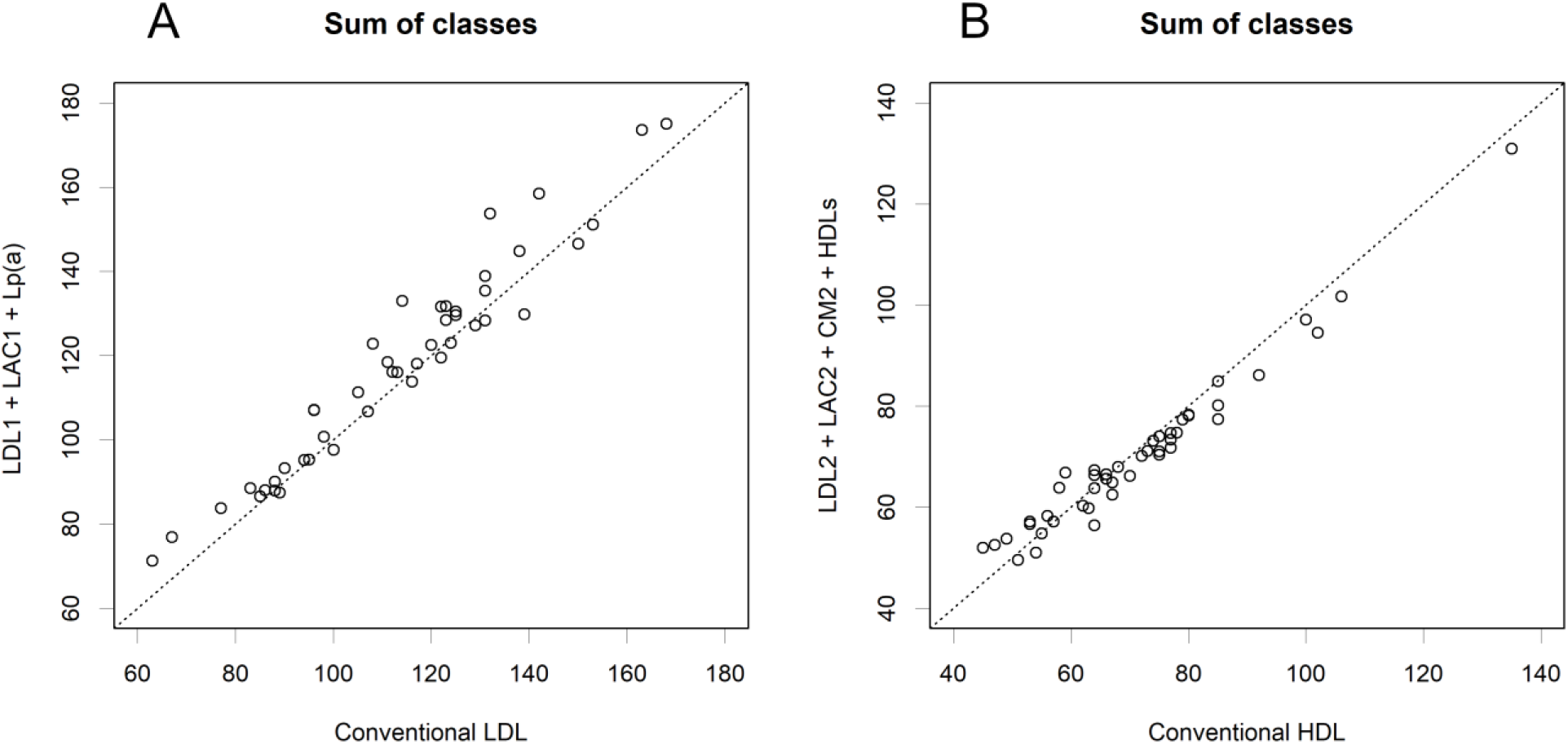
Estimation of the sets of classes that are measured by a conventional method. If the conventional methods extract most of the cholesterol from particular classes, these would present the sum of the quantities of the classes. The combinations presented here were the ones with the highest correlation and the closest amounts.

Thus, the perception of lipoproteins should be updated. They are classes of protein-rich particles, each of which has specific functions. Only limited apoproteins were studied here, but the attributes of other apoproteins are important for their functions. Hence, the classes need to be studied extensively, which will provide a deeper understanding of the pathophysiology. The levels of the classes varied among volunteers and were lognormally distributed. Some classes are narrowly controlled and are good candidates for indicators of diseases. Conventional enzymatic methods measure mixtures of multiple classes. Because the sums of lognormally distributed numbers are not informative (Fig. S12), the data do not provide a proper diagnostic criterion. This could be the reason why levels of conventional LDL did not indicate the prognosis of patience ^19^. However, HPLC is not suitable for large numbers of samples. Therefore, simpler methods for measuring specific classes are required. In particular, the targets of enzymatic methods should be completely refined. According to the lognormal distribution characteristics, independent clinical treatments, such as antilipidemic drugs, nutritional therapy, and ergotherapy, may synergistically change the levels of specific classes.

## Data Availability

All the data is available at Figshare https://doi.org/10.6084/m9.figshare.14247119.v1

## Acknowledgements

We would like to thank Editage (www.editage.com) for English language editing.

## Supplementary Materials

Supplementally figures and tables are in Figshare ^20^.

### Materials and Methods

Blood samples: Samples were collected after obtaining informed consent from all volunteers and approval from the ethics committee of Akita Cerebrospinal and Cardiovascular Center (ID. 19-21). All samples were anonymized prior to analysis. No postmeal time was specified for blood collection. The age of the volunteers is shown in Figure S1.

The collected blood was sent to a clinical laboratory (SRL Inc., Tokyo, Japan). The serum was separated, and HLD, LDL, and total cholesterol were measured using conventional enzymatic methods. The serum was sent to Skylight Biotech Inc. (Akita, Japan) for further analysis using gel filtration HPLC. Forty-four healthy volunteer samples were subjected to analytical HPLC, and TG and cholesterol were monitored sequentially. Furthermore, 11 samples (6 healthy and 5 hyperlipidemic) were subjected to preparative HPLC and then fractionated.

The HPLC monitoring data (TG and cholesterol) were analysed parametrically ^8^. This method is a parsimonious way of performing curve fit to maintain the falsifiability of the model using the minimum number of classes assumed ^21^. With many estimated classes, the fitting process will become easier; however,the assumed classes must be verified by reality. Too many assumptions make this verification difficult. The size and range of a class are presented using the position μ and scale σ of the normal distribution. We assumed that a class would contain TG and cholesterol at a certain constant rate regardless of the size differences within the class., with The amount of each TG and cholesterol presented by using another parameter. This assumption was verified through the curve-fitting process.

The standard values of TG and cholesterol in each class were estimated from the full data set using a trimmed mean (0.2). Their 95% range was estimated using the median absolute deviation (MAD): the upper and lower limits of the 44 healthy samples were estimated as trimmed mean to two MADs. The standard values of the position or scale parameters of the classes, which were varied, were estimated from the trimmed mean of μ or σ^2^ found in each sample.

In preparative HPLC, the elution was periodically fractionated. UV was also monitored, and the protein amounts of CM and VLDL were estimated under the assumption that any protein absorbs a fixed amount of UV per weight. Each fraction was subjected to 5 %–20% SDS-PAGE, and the proteins were detected using silver staining. Some protein bands were identified using MALDI-TOF MS (Genomine Inc., Kyungbuk, Korea). In addition, specific proteins were confirmed by western blotting after transfer to PVDF membranes. The antibodies used were as follows: anti-apoB antibody (A-6), sc-393636 AF488; apoA-I antibody (B-10), sc-376818 AF647 (Santa Cruz Biotechnology Inc., Texas, USA); anti-Lipoprotein a antibody, ab27631, (Abcam plc., London, UK). Chemiluminescence of the antibodies and silver-stained gel bands were measured using an Amersham Typhoon Scanner (Cytiva).

## Notes

### Competing Interest Statement

The authors have declared no competing interest.

### Clinical Trial

NA

### Funding Statement

This study is supported by the scholarship donation of Akita Prefectural University.

### Author Declarations

The ethics committee of Akita Cerebrospinal and Cardiovascular Center (ID. 19-21, 2018/5/7)

### Summary of Updates

I am so sorry but I mistakenly added "1" in the title. This was removed.

